# Improving healthcare team harmony through collaborative team reflection and mindfulness

**DOI:** 10.1101/2023.07.25.23293060

**Authors:** Matthew JY Kang, Ar Kar Aung, Jo Gibbs, Harry Gibbs

## Abstract

Hospital wards, staffed by the multidisciplinary team, are complex environments where teamwork, communication and psychological safety is essential for coordinated care delivery, yet are faced with challenges such as staffing changes and complex care needs. However, there is little literature on interventions to assist staff connect as a team. We evaluated a brief daily group based on team reflection and mindfulness aimed at a multidisciplinary general medicine team, using measures of team functioning (effectiveness, communication, and psychological safety).

We found that participants reported significant improvement in the meeting’s effectiveness (*U=*184, p=0.013), team morale (*U=*123, p<0.001), and focus (*U=*183, p<0.001) after the program’s commencement. Furthermore, participants who attended the program for at least a week reported they felt more psychologically safe (*U=*116, p=0.032). We also found significant positive correlation between measures of team functioning and the number of sessions they attended the program (effectiveness of the interdisciplinary meeting r=0.509, p<0.001; team’s communication and functioning (r=0.509, p<0.001). The post-intervention focus group highlighted the program helped build relationships within the team, improve psychological safety, and subsequently shifted the team’s behaviour to be more supportive of the overall team. Our program improved the functioning of a multidisciplinary healthcare team, with the overall aim to deliver better patient care.

## Background

Hospital wards are dynamic environments where effective communication and coordination among multidisciplinary staff are crucial for quality care delivery (Rosen et al., 2018).

Multidisciplinary teams, and interdisciplinary rounds, offer numerous benefits in terms of coordinated care and clinical problem-solving (Henkin et al., 2016; Ryan et al., 2017). Although there is research in ways to operationalise multidisciplinary rounds, there is a lack of evidence regarding strategies to enhance team communication and functioning in clinical settings.

Recently, we evaluated the “Collective Pause” program, a modified group-based mindfulness-based intervention (MBI) aimed at healthcare teams. (Kang et al., 2020) We found that Collective Pause was positively received by staff, resulting in improved attention during the meetings. In addition to the individual benefits in feeling more mindful, the staff noted that the group MBI program helped feel more connected to each other as a team, which is linked with better collaboration and improved conflict management (Good et al., 2016). We believed this was an important benefit of the program that needed more development and investigation.

The project aimed to determine whether a novel teambuilding program based on collaborative team reflection and mindfulness, designed to be brief, regular, and conducted in order to promote communication and build relationships within the multidisciplinary team in healthcare setting, can enhance team functioning and effectiveness.

## Methods

### Setting

This program was delivered to the staff of the multidisciplinary general medicine team at Alfred Health. The disciplines included nursing, medical, administration, occupational therapy, physiotherapy, pharmacy, social work, and speech pathology. Participant numbers ranged from 20 to 30 each session, with all staff on shift invited to attend, with a couple staff nominated to be available for urgent patient care. The program took place in the staff nursing station immediately prior to their daily morning interdisciplinary round (Mon-Fri), for six-weeks, and was optional for staff.

### The Program

Each session of the program involved three phases, guided by a trained facilitator who was not a team member. We started with each participant sharing their name and their energy and emotional state with the group. The session ended with a mindful pause, where each participant was guided through breath modulation and self-affirmations to set a positive intention for the rest of the day. Each session lasted no more than 10 minutes.

The program was supported by senior ward leadership. Participants provided written informed consent, and there was no reimbursement for their involvement. Variations in participants occurred naturally due to staff rotation and rostering.

### Quantitative data

We collected de-identified information about staff members including age and discipline one week prior (baseline) to the commencement of the program. Participants were asked to complete weekly surveys where they rated their agreement or disagreement on a Likert scale (ranging from 1 for strongly disagree, 7 for strongly agree) based on statements relating to four domains:

Domain 1 - Four questions to measure the effectiveness of the meeting based on a four-question survey that we developed from a previous study that examined the perceived benefit of interdisciplinary rounds “My team was ‘on the same page” about the patient’s care including discharge planning’ (Walton et al., 2019), and Domain 2 - Four questions to explore the participant’s overall perception of the team’s psychological safety during the week, using the participatory safety subscale of the Team Climate Inventory (based on statements (i.e. “We have a ‘we are in it together’ attitude”) (Kivimaki and Elovainio, 1999).

Domain 3 - One question to ascertain morale of the team (“The team morale was high “) (Woods and Almvik, 2002),

Domain 4 - One question to ascertain individual focus (“I was able to maintain focus during the week”),

We summed the questionnaire scores of the participants’ perception of the meeting effectiveness (scores ranging from 0-28) and team’s functioning overall (scores ranging from 0-28). As the data was not normally distributed, we used Mann-Whitney U tests (U statistic, p-value), to compare the measures of team functioning and meeting effectiveness between the teams prior to and after the program commencement. Spearman’s correlation was used investigate the accumulative impact of the program (number of sessions) on the above measures. Given this was a naturalistic study with varying membership of participants in each session, a paired statistical analysis was not undertaken.

### Qualitative data

We organised a voluntary focus group, and invited staff to represent each discipline and seniority to participate and share their experience of the program. The focus group was audiotaped and transcribed, which underwent thematic analysis by MK following Braun and Clarke’s six-stage method involving familiarisation with data, generation and collation of codes into themes, reviewing themes in relation to the dataset and defining and reporting on the hierarchy.

This project was approved by the Alfred Health Ethics Committee (Project Number #501/22). Prior to project commencement, participants were given an information sheet about avenues to seeking professional support for their health.

## Results

Forty-six unique participants completed the survey, with the mean age of 32.2 years (SD 13.0) and 71% females. There was representation from medical (n=20), nursing (n=5), pharmacy (n=5), physiotherapy (n=3), dietetics (n=3), social work (n=2), occupational therapy (n=2), administration (n=2), neuropsychology (n=1), and other allied health (n=1). Two people did not complete details about their occupation nor sex.

Following commencement of the program, there was significant increase in the team’s perception of the meeting’s effectiveness (*U=*184, p=0.013), team morale (*U=*123, p<0.001), and focus (*U=*183, p<0.001), whilst a trend for the team’s psychological safety (*U*=194, p=0.090). When we examined subjects who had participated in the program for at least a week (5 or more sessions), the team’s perception about their psychological safety significantly increased (*U=*116, p=0.032), whilst the other measures (perception of meeting effectiveness, team morale and focus) remained significantly increased as well.

**Table 1.** Measures of team functioning comparing baseline vs final measures of all participants who attended at least one sessions vs one week (five sessions)

|  | Baseline (n=17)* | Attended at least one session (n=40**) | Attended at least one week (n=28**) |
| --- | --- | --- | --- |
| Psychological safety | 21.0 (IQR 4.0) | 23.5 (IQR 4.0)<br>$U=194$ ($p=0.090$) | 24.0 (IQR 5.25)<br>$U=116$ ($p=0.032$) |
| Meeting effectiveness | 16.0 (IQR 2.5) | 18.0 (IQR 2.25)<br>$U=184$ ($p=0.013$) | 18.5 (IQR 3.0)<br>$U=102$ ($p=0.003$) |
| Team morale | 4.0 (IQR 1.0) | 6.0 (IQR 1.25)<br>$U=123$ ($p<0.001$) | 6.0 (IQR 2.0)<br>$U=59$ ($p<0.001$) |
| Individual focus | 4.0 (IQR 1.0) | 6.0 (IQR 1.0)<br>$U=183$ ($p<0.001$) | 6.0 (IQR 1.0)<br>$U=101$ ($p<0.001$) |

Expressed as median (IQR) IQR = interquartile range, $U$ = Mann-Whitney U Test, $p$ = p-value
\*3 missing data for psychological safety, 2 for team morale and 1 for meeting effectiveness
\*\*1 missing data for individual focus

There was significant positive correlation between the number of sessions participants attended and the effectiveness of the meeting (r=0.435, p=0.003; Figure 1A), team’s psychological safety (r=0.509, p<0.001; Figure 1B), team morale (r=0.724, p<0.001) and individual mindfulness (r=0.703, p<0.001).

**Figures 1A.**
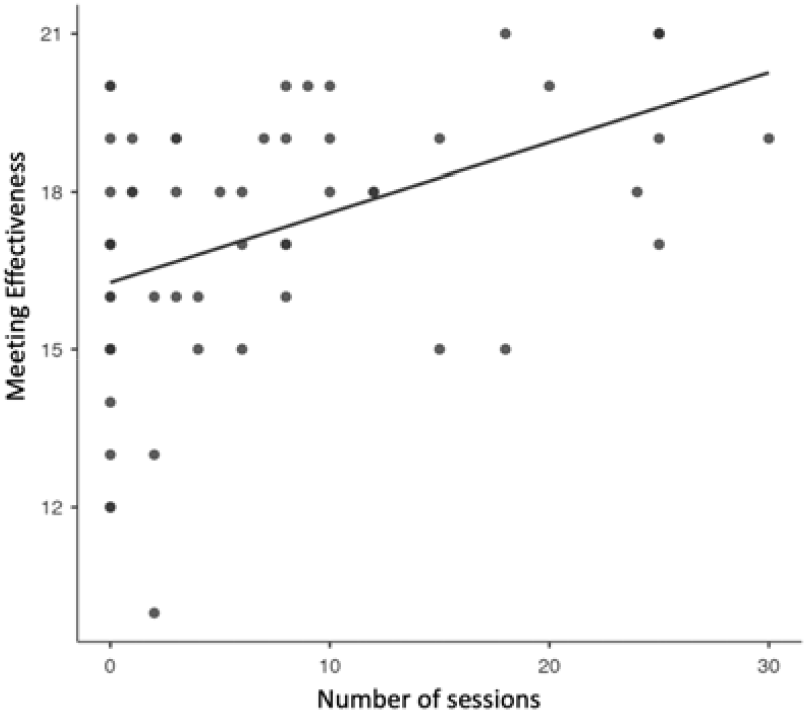
correlation between number of program sessions and meeting effectiveness.

**Figures 1B.**
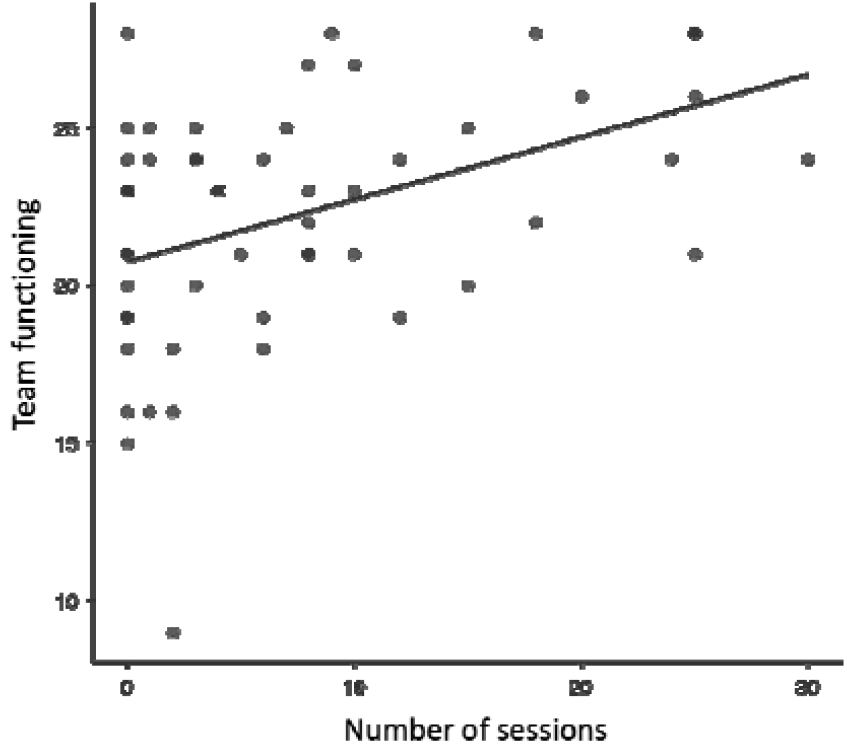
Correlation between number of program sessions and the team’s psychological safety.

**Figure 2.**
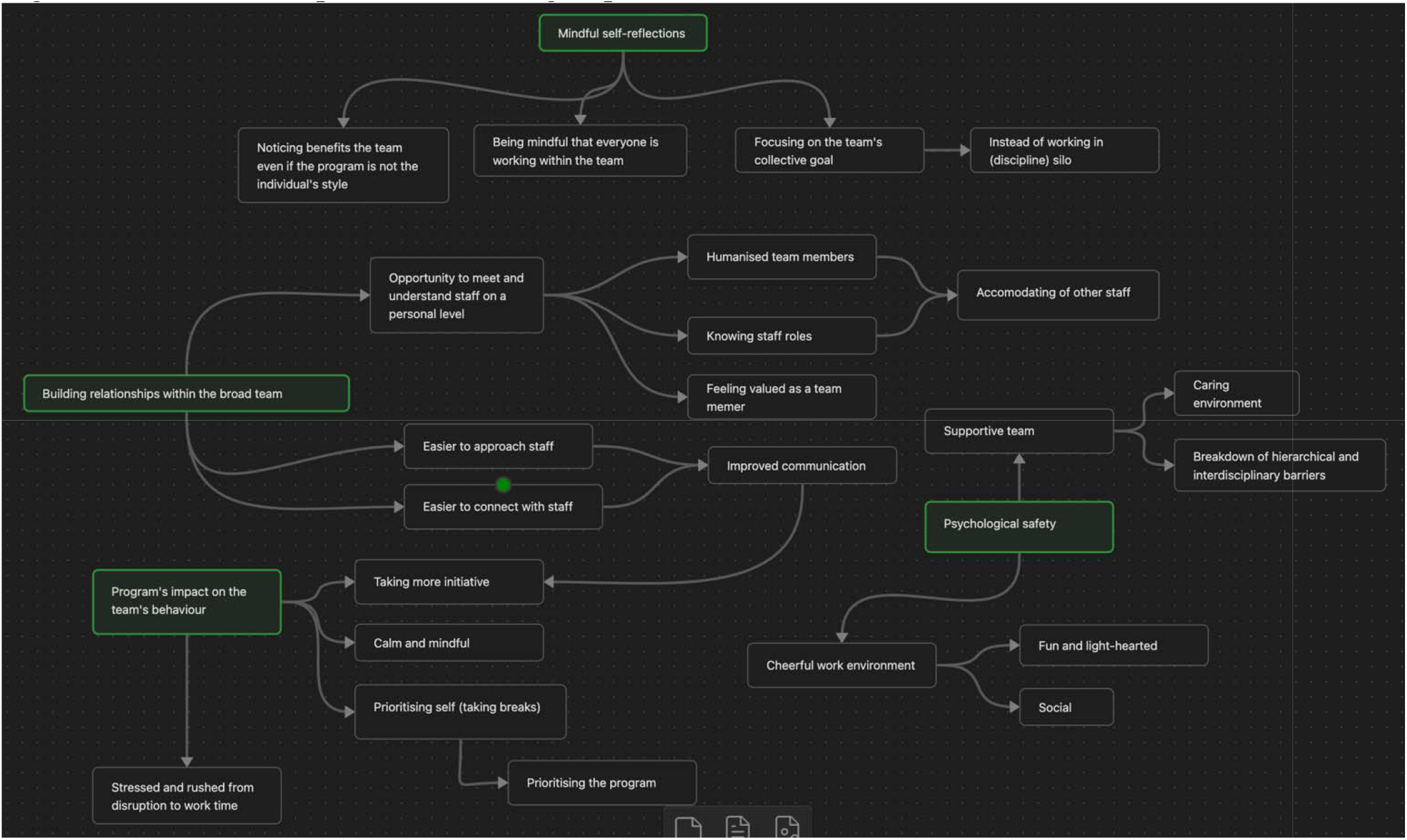
Thematic map from the focus group.

Nine staff members participated in the focus group. Four broad themes emerged from the focus group, which were mindful self-reflections, building relationships within the broad team, program’s impact on the team’s behaviour and psychological safety.

### Building relationships within the broad team

The participants felt the program allowed them an opportunity to meet members of the multidisciplinary team. Moreover, the the program allowed the staff to have an opportunity to reflect together and be vulnerable about their current wellbeing state. The participants expressed that this humanised each team member, which led them to being more supportive of each other and their respective workloads. The simple gesture of knowing each other’s names and roles led to staff feeling more valued as a member of the team.

As a result of the relationship building, the participants noted it was easier to approach and connect staff members. This led to improved communications within the team, and also more usage of staff’s expertise, which participants reflected was the result of the staff being better familiar with each other’s roles.

### Psychological safety

Participants highlighted the fun and light-hearted nature of the program, which made the team environment more social. They also noted that the program, which senior staff members supported to promote staff wellbeing, fostered a caring environment within the team, which was a relief especially for more junior staff who were new to the team. They also noted that the program’s activities helped breakdown hierarchical barriers through its light-heartedness and familiarity with team members. Participants noted that this made it easier to approach senior staff members, including medical consultants, to ask questions or raise issues about patient care.

### Program’s impact on the team’s behaviour

Some participants noted that as a result of the improved communication amongst the team, this led to them taking more initiative in the workplace as they had a better sense of whether a particular staff needed more assistance.

Moreover, the participants reflected the program made them feel calmer and more mindful during work, including of their own wellbeing. This led them to prioritise their breaks at work, which previously may have been neglected due to the busy workload.

Notably, the program made some participants feel more stressed at times, as it meant less time was available for their clinical duties. Despite this, the participants noted they prioritised the program in their day when they could, though wondered whether a weekly option or sessions in the afternoon would have allowed them to plan their day differently.

### Mindful self-reflections

The focus group highlighted that the program helped them become more mindful that they were part of a team with a collective goal, whereas prior to the program, they felt like they were working in a silo within their discipline.

**Table 2.** Select subthemes and quotes from the focus group.

| Theme | Quotes |
| --- | --- |
| Having fun in a light-hearted environment | “I don't think it mattered what the activity was, whether it was squats, star jumps, I think it was the fact of getting us all together, having fun, nobody cared how well we squatted.”<br>“(the program) brought a life and fun... it's that nice juxtaposition between the heaviness on the ward, and starting with the lightness and clarity before launching into the day. “ |
| Building relationships | “I’d say from my perspective, it enabled the initial introduction to people you may not have known.” |
| Opportunity to meet and understand staff on a personal level | “I think it helped humanise the team members”<br>“When you start (the program), it makes it easier to just on a personal level get to know them, and also made communication easier on the ward.” |
| Psychological safety | “It made me feel psychologically safe, the environment and vibe of the ward was like “we are focused on mental wellbeing”, we are talking about how we are going as people and feel appreciated about you as a person, and connect with another person as a person, not just another coworker.” |

## Discussions

Promoting connectedness in the multidisciplinary team is crucial for fostering effective collaboration and communication among staff, thereby enhancing patient care outcomes. We found that our program led to significant improvements in the team’s perception of their psychological safety and communication generally, as well as during the interdisciplinary meeting.

Previous literature examining teams, including those outside of healthcare, have found that psychological safety was the most important attribute for teams to operate most effectively (Grailey et al., 2021). Psychological safety enables the staff to be their “true selves” and minimise medical errors. The improvements in psychological safety from the program are supported by the qualitative data, by breaking down hierarchical barriers, creating a supportive environment that prioritises each staff’s wellbeing, and enabling the team to be more social and connected.

Our finding that the program enabled participants to become more mindful and reflective of their team working towards a collective goal rather than individual silos is in line with our previous study that the program improved individual’s ability to decenter, defined as the ability to be open, curious and accept the present experience (Kang et al., 2020; Lau et al., 2006). In the busy environment of an acute medical ward, our program reminded staff of the importance of working as a team, rather than feeling alone in the system, which has been associated with increased burnout (Karcz et al., 2022).

We also found the program helped foster a psychologically safe workplace, which is linked with safe care delivery and quality improvement (Grailey et al., 2021). The program also facilitated increased communication within the team, which is crucial in an environment where there is constant changes in staff personnel. Furthermore, breaking down hierarchical and interdisciplinary barriers further helped improve communication within the team.

Despite the overall positive feedback from the participants, fitting even our brief program had costs. Asking staff of more of their time despite their workload may have been challenging on particularly stressful days. We also need to acknowledge that each staff have their own way of communication, maintaining calm and self-reflections, which may not be in line with the program’s activities. Nonetheless, even the staff who were not individually benefitting from the program observed that there were improvements in the overall team’s communication and relationship, which highlights the net positive effect.

The limitations of this study include the lack of a control group and selection bias. The generalisability may be limited as this is a single-centre study, and the internal validity also needs to be confirmed in an alternative setting (e.g. a non-medical ward). Furthermore, as outcomes for patient care are not measured, we have yet to demonstrate that the observed improvements in team communication and relationships indeed translate to meaningful improvements in patient care.. However, objective measures of team communication and functioning by an independent observer add further rigour to our findings.

In conclusion, our program enabled meaningful connections between the multidisciplinary healthcare team, promoting psychological safety and better communication. As the healthcare system is a dynamic environment where staff personnel change constantly, fostering a positive team environment is a challenge. Our simple program demonstrates the value of spending 10-minutes a day to connect the team based on mindfulness and wellbeing principles, to foster trust and communication within the team, ultimately improving team functioning for better patient care.

## Data Availability

All data produced in the present study are available upon reasonable request to the authors

